# A special case of COVID-19 with long duration of viral shedding for 49 days

**DOI:** 10.1101/2020.03.22.20040071

**Authors:** Li Tan, Xia Kang, Bo Zhang, Shangen Zheng, Bo Liu, Tiantian Yu, Fan Yang, Qiongshu Wang, Hongming Miao

## Abstract

Prolonged viral shedding is associated with severe status and poor prognosis of COVID-19 patients. Unexpectedly, here we report a non-severe patient with the longest duration of viral shedding. According to the investigation on the clinical and epidemiological information of this case, we concluded that this type of virus might have a low toxicity and transmissibility, but have a prolonged infective ability and was hardly to be eliminated in the body with regular therapy. However, infusion of plasma from recovered patients showed high efficiency in elimination of this virus. Our findings might shed light on the management of COVID-19.

## INTRODUCTION

Coronavirus disease-19 (COVID-19), caused by severe acute respiratory syndrome coronavirus 2 (SARS-CoV-2), has become a threat to global health^1-3^. The clinical spectrum in COVID-19 patients presents diversely^4-5^. Viral shedding is a critical indicator for prognosis, and prolonged viral shedding always predicts a poor outcome^5-6^. In this report, however, we describe a special case of a family cluster with a mild type of COVID-19. In one family member, the duration from illness onset has persisted for over 49 days till now, which is the longest duration of viral shedding in symptomatic patients as far as we know. Nevertheless, this case shows a mild infectivity and a better status comparing with most cases. Fortunately, treatment with plasma from recovered COVID-19 patients efficiently cleared out the virus infection. This report describes the detailed epidemiologic and clinical information of this special patient and his close contacts, which might provide rationale for reasonable treatment, risk-stratification, strategy of community healthcare management and policymaking for hierarchical medication.

## METHODS

### Disease classification of COVID-19

All cases were diagnosed and classified according to the New Coronavirus Pneumonia Diagnosis Program (5th edition)^7^ published by the National Health Commission of China. Clinical manifestations consist of four categories, mild, moderate, severe and critical. The mild clinical symptoms were mild with no pulmonary inflammation on imaging or without symptoms of respiratory infections. The moderate is the overwhelming majority, showing symptoms of respiratory infections such as fever, cough, and sputum, and pulmonary inflammation on imaging; when symptoms of dyspnea appear, including any of the following: shortness of breath, RR ≥ 30bpm, blood oxygen saturation ≤ 93% (at rest), PaO_2_ / FiO_2_ ≤ 300 mmHg, or pulmonary inflammation that progresses significantly within 24 to 48 hours> 50%, it was classified as severe; respiratory failure, shock, and organ failures that require intensive care were critically ill.

### Patients information

In this study, all cases were taken from one of the designated hospital for the COVID-19 by local authority in Wuhan, a city of Hubei province in China. This study was approved by the Ethics Committee of the hospital. All subjects signed informed consent forms at admission to hospital. 130 patients including the discharged and dead in our hospital between January 14, 2020 and March 19, 2020 were investigated and their clinical indexes were used as references of a hospitalized patient (Case 1), who would be introduced in detail in this study. There were 96 cases of moderate type, 19 cases of severe type and 15 cases of critically ill type (all died). Another patient Case 2, the close relative of Case 1, was also followed. All patients were confirmed by viral detections using quantitative RT-PCR^8^, which ruled out infection by other respiratory viruses such as influenza virus A, influenza virus B, coxsackie virus, respiratory syncytial virus, parainfluenza virus and enterovirus by the same time.

### Data collection

In this study, the basic information, clinical symptoms, complete blood count, coagulation profile, and serum biochemical test (including renal and liver function, creatine kinase, lactate dehydrogenase, and electrolytes) and disease outcome of all included patients were collected. The epidemiological data of the investigated patient Case 1 was also collected.

### Statistical methods

In this study, GraphPad 6.0 software was used for data statistics and mapping. All the data were displayed descriptively.

## RESULTS

### Case 1

A middle-aged man visited our hospital to require SARS-CoV-2 tests on February 8, 2020. The patient stated that he had got an intermittent fever and continued for around 1 week since January 25 without other typical symptoms of COVID-19, including chills, stuffiness, headache, dry cough, pharyngalgia, chest pain, shortness of breath or diarrhea. The highest body temperature was 38.1 °C. After taking antipyretics, Chinese traditional medicine and anti-viral medicine by himself, the temperature decreased to normal levels in one week. Since his close relative was confirmed to be infected with COVID-19 one day before, he asked for further examinations. On admission, the patient reported no subjective symptoms. Physical examination revealed a body temperature of 36.2 °C, respiratory rate of 14 breaths per minute, pulse of 88 beats per minute and blood pressure of 112/68 mmHg. Lung auscultation revealed no rhonchi and crackles. Chest CT scan showed that infective signs were identified in superior lobe of right lung and inferior lobes of bilateral lungs. Laboratory tests showed the number of white blood cells, number of lymphocytes, percentage of lymphocytes and percentage of neutrophils were in normal levels (Table 1). Nasopharyngeal swab specimens were obtained to test for influenza A and B and the results were negative. After admission, the patient received anti-viral therapies and supportive care.

**Table 1.** Basic and clinical information of two COVID-19 patients on admission

| Items | Case 1 | Case 2 |
| --- | --- | --- |
| **Demographics & clinical characteristics** |  |  |
| Age range (years) | 40-50 | 70-80 |
| Sex | male | female |
| ABO | AB | NA |
| Smoking | N | N |
| Comorbidity | N | Rheumatoid arthritis |
| Respiratory rate (on admission) | 14 | 20 |
| Pulse (on admission) | 88 | 82 |
| Systolic blood pressure (on admission) | 112 | 138 |
| Diastolic blood pressure (on admission) | 68 | 69 |
| Fever | Y | Y |
| Cough | N | Y |
| Diarrhoea | N | N |
| Nausea | N | N |
| Chest pain | N | N |
| Disease severity status | Moderate | Moderate |
| Time from illness onset to hospital admission (days) | 15 | 10 |
| **Laboratory findings (on admission)** |  |  |
| Red blood cell count ($\times 10^{12}/L$) | 4.56 | NA |
| Hemoglobin (g/L) | 137 | NA |
| Platelet count ($\times 10^9/L$) | 222 | NA |
| White blood cell count ($\times 10^9/L$) | 6.6 | 6.95 |
| Lymphocyte count ($\times 10^9/L$) | 2.19 | 1.21 |
| % Lymphocyte | 33.4 | 17.40 |
| Neutrophil count ($\times 10^9/L$) | 3.61 | 4.91 |
| % Neutrophil | 55.1 | 70.70 |
| Monocyte count ($\times 10^9/L$) | 0.68 | NA |
| % Monocyte | 10.4 | NA |
| Albumin (g/L) | 41.9 | 38.4 |
| ALT (U/L) | 24 | NA |
| Creatinine ($\mu\text{mol}/L$) | 79 | 89.8 |
| Uric acid | 232 | NA |
| Prothrombin time | 11.8 | NA |
| D-dimer | 90 | NA |
| IL-6 (pg/mL) | 2.3 | NA |
| **Imaging features (on admission)** |  |  |
| Bilateral pulmonary infiltration | Y | Y |

N, No; Y, Yes; NA = Not available

On days 2 through 5 of hospitalization, the patient presented with intermittent low fever, with the highest body temperature under 37.5°C and other vital signs stable (Fig. 1A). A nucleic acid amplification test (NAPT) for COVID-19 was positive (Fig. 1A and 2D). Re-examination of Chest CT scan showed obvious resorption of the infection lesions of bilateral lungs. From hospital days 6 till now, the overall status of Case 1 remained stable. The patient presented with normal levels of vital signs including body temperature (Fig. 1A). Nevertheless, COVID-19 testing on specimens collected from oropharyngeal swabs on illness days 17, 22, 26, 30, 34, 39, 43 and 49 were positive and on illness day 47 was negative (Fig. 1A). The detailed information on primary laboratory tests can be found in Fig. S1. Noteworthily, the levels of blood lymphocytes, interleukin-6 and procalcitonin have been recognized as indicators for disease severity and prognosis of COVID-19 patients by us and others^5,9^. These three indexes were all normal and stable in this patient (Fig. 2A-C), while the viral load persisted in a high level as the severe or critical ill patients did (Fig. 2D). Notably, IgG antibody against SARS-CoV-2 showed positive on Feb. 20 and Mar. 14, while IgM antibody tests remained negative.

**Fig. 1.**
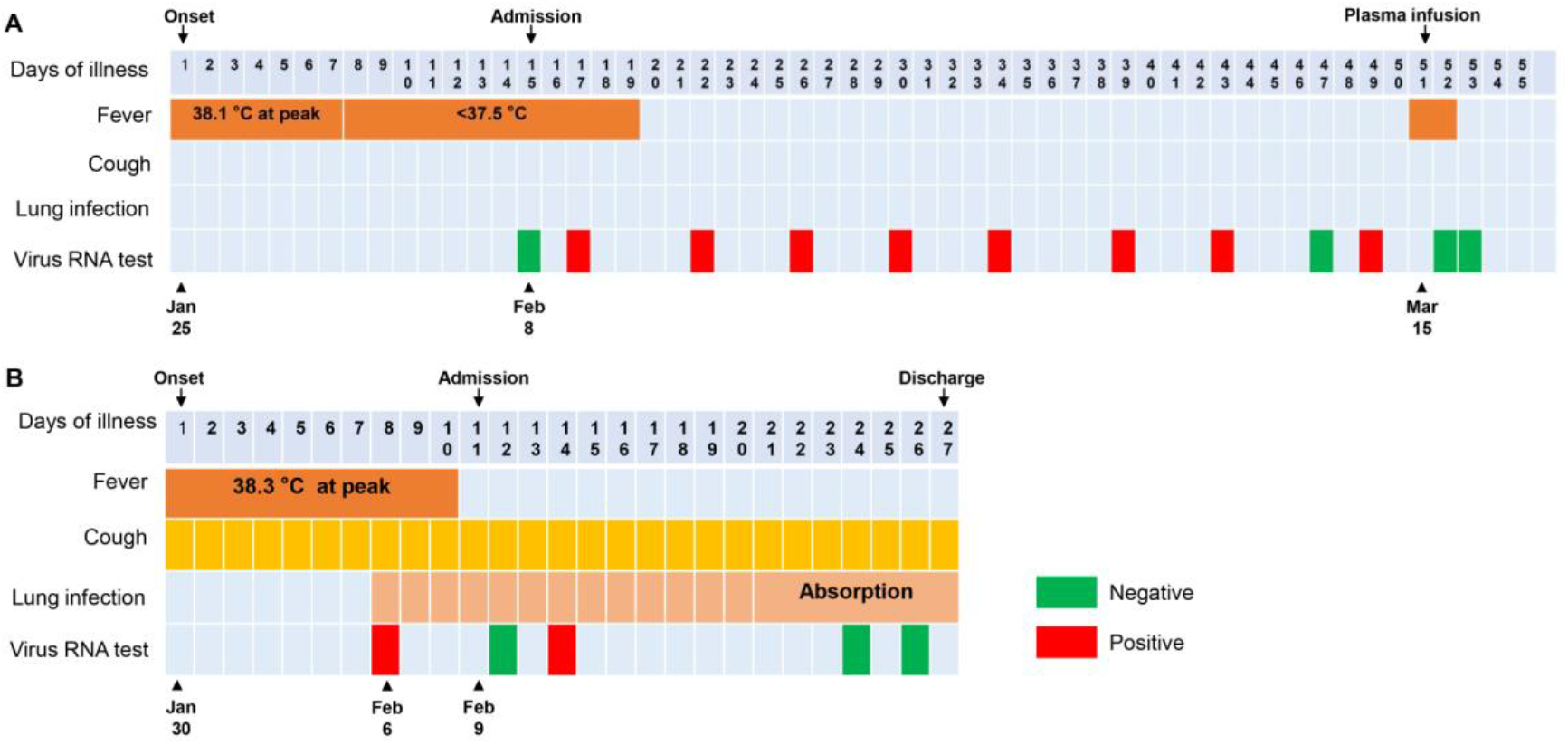
Dynamic assessment of symptoms, body temperatures and viral infection status of Case 1 (**A**) and Case 2 (**B**) from January 25 to March 19, 2020.

**Fig. 2.**
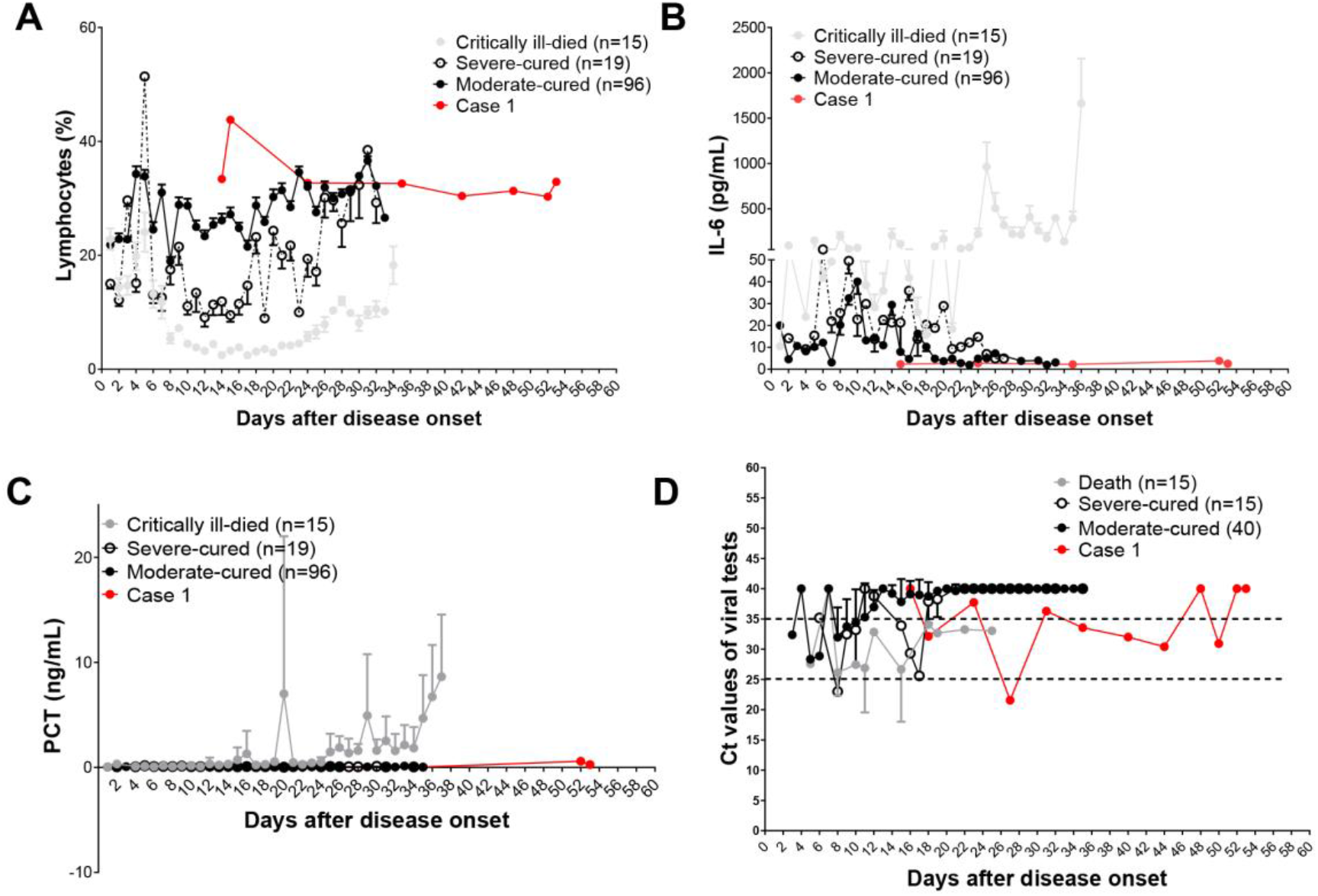
Dynamic changes of lymphocyte percentage (A), IL-6 levels (B) and PCT levels (C) in peripheral blood and Ct values of viral tests (D) in the moderate-cured, severe-cured and critically ill patients and Case 1 with COVID-19. The parameters of cured or dead patients were used as references of Case 1. Severe-cured, severe type of patients with a cured outcome; Moderate-cured, Moderate type of cases with a cured outcome; IL-6, interleukin-6; PCT, procalcitonin

Given the prolonged infection, this patient received plasma infusion treatment. The plasma was collected from recovered patients with COVID-19. On Mar. 15, Case 1 received 400 ml of plasma. The patient got fever in 4 hours after infusion, with the body temperature up to 38.9 °C, which we considered as a blood transfusion reaction. As expected, the body temperature became normal the next day. Notably, the viral tests of SARS-CoV-2 via oropharyngeal swabs on Mar. 16 and Mar. 17 turned into negative.

Epidemiological survey revealed that besides his relative, the patient closely contacted with another five people before admission. One contact also got fever for a short term but the test for SARS-CoV-2 showed negative. The other four contacts without any symptoms were also negative in SARS-CoV-2 tests.

### Case 2

Case 2 is a close relative of Case 1. This elderly woman visited our hospital on February 9, 2020. She complained an intermittent fever combined with occasional dry cough for around 10 days. Before visit, the highest body temperature was 38.3 °C. On February 6, Chest CT showed bilateral lung infective signs in this patient. Laboratory tests revealed an elevation of neutrophil percentage (82.3%) and C reactive protein level (CRP) (61 mg/L). The patient was prescribed anti-viral medicines and Chinese traditional medicines. However, the fever was not absolutely controlled after receiving these treatments for 3 days. Notably, this patient has a history of rheumatoid arthritis (RA) with long term use of methylprednisolone (4 mg, oral, 1 qd) (Table 1 and Fig. 1B).

After admission, physical examination showed a body temperature of 36.0 °C, respiratory rate of 20 breaths per minute, pulse of 82 beats per minute and blood pressure of 138/69 mmHg. Lung auscultation revealed no rhonchi and crackles. Re-examination of Chest CT scan on admission revealed exudative lesions in bilateral lungs and the result on hospital days 5 became worse. However, on hospital days 11, Chest CT scan revealed the lung infection attenuated significantly. For laboratory tests, the results on hospital days 2 revealed that the percentage of neutrophils (70.70%) and CRP level (9.81 mg/L) dramatically decreased comparing with the results on 5 days before. SARS-CoV-2 tests presented positive on specimens collected from oropharyngeal swabs on hospital days 4, but it turned into negative on hospital days 14 and 16. Then the patient was permitted to discharge from hospital (Table 1 and Fig. 1B).

## DISCUSSION

It was reported that the median duration of viral shedding was 20.0 days from disease onset and the longest was 37 days^5^. There were no significant differences between severe patients (19 days) and critical patients (24 days)^5^. In our report, the duration of viral shedding from illness onset in Case 1 has persisted for 49 days, which has been the longest in ever reported in symptomatic patients. Previous studies indicated the level and duration of viral shedding is a critical indicator to access the risk of transmission and to guide the isolation of patients as well as predicting the prognosis^5-6^. In viral infection, prolonged viral shedding was associated with inferior outcome^6^. Interestingly, contrary to the conclusions above, we here reported one of the non-severe cases has the longest duration of viral shedding. The Case 1 only got moderate fever initially and the body temperature rapidly decreased into normal levels without any respiratory failure. Although RT-PCR analysis showed virus was not eliminated, the symptoms and signs have been largely stable after admission. Notably, besides Case 1, all close contacts revealed mild responses to this virus. Case 2, an elderly woman, got fever for about 10 days while the highest body temperature did not exceed 38.5°C, although the lung infection once got worsen, the progression was controlled in several days. Given that Case 2 has a history of long-term use of glucocorticoids, which is the high-risk factor for severe infection and progression, overall, the severity of Case 2 was much optimistic than average status in her age group, since it was reported elder people infected SARS-CoV-2 have much worse prognosis^10^. Furthermore, though his another relative also got fever at the time of visit, the viral test was negative, indicating the virus may be promptly cleared off or this contact was not infected. While genetic mutations may be considered to contribute to the mild signs in this family cluster case, one more important thing is other four close contacts who do not have any blood relationship with Case 1 even were not infected after contact. Recently, one study found ABO blood group is a new risk factor for severity of patients with COVID-19. People with blood group A have a higher risk and the patients with blood group O have a lower risk than other groups^11^. However, the blood group of Case 1 is group AB (Table 1), thus excluding the possibility that blood group of Case 1 may influence the severity of illness. These clues together indicated that the SARS-CoV-2 virus in Case 1 and Case 2 may be a mild subtype, both to young and to old people. Tang et al. investigated the molecular divergence of SARS-CoV-2, showing the virus evolved into two major types (namely L subtype and S subtype)^12^. Of which L type is more prevalent consisting of approximately 70% in all patients and S type is the ancestral version. In addition, L type has a high frequency in patients and presents more likely to spread comparing with S type^12^. Currently, few studies focused on identifying the clinical features between these two subtypes, thus we cannot assure that Case 1-associated virus belongs to S type, mutated L type or a new subtype. Since the onset time of Case 1 revealed this mild subtype already exists at the early stage of outbreak when L type occupied dominant position, we cannot exclude an original new subtype that was not identified. Because this potential subtype shows a quite low toxicity, weak transmissibility and people infected with this type showed an affirmative outcome, it is worthy for us to further analyze the mRNA sequence of the virus type isolated from Case 1, which will help us to distinguish potential mild patients. It is beneficial to distribute the limited medical sources and to guide the community healthcare management in the context of pandemic.

Due to the limited capacity of nucleic acid assays, identifying the reliable indicators to distinguish the severity of patients in early stage is helpful to quickly screen out the patients who needed to be hospitalized or to stay at home. The status of lymphocytes and duration of viral shedding are two most common indicators in predicting prognosis of COVID-19 patients. However, it is still unclear which one of them is more reliable and more efficient. In the present study, IgG antibody can be detected in Case 1 from Feb. 20, indicating Case 1 can initiate immune responses to viral infection. However, the mRNA level of virus in Case 1 showed a similar trend to that of death cases. Taken together, these findings indicated that immune indicators could be more sensitive and reliable than viral shedding in reflecting the overall status of patients and predicting the prognosis. Actually, a recent study found that recruitment of T cells and the presence of IgG and IgM antibodies can be found before the resolution of symptoms in COVID-19 patients^13^. Therefore, we strongly suggest that immune indicators, especially the level of lymphocytes, could be a better prognostic indicator than duration of viral shedding, especially for patients in early stage.

We next would like to find any other signs that may also be potential prognostic factors besides the level of lymphocytes in early stage of illness. It was reported that the median duration of fever in COVID-19 patients was about 12.0 days^5^. Obviously, the duration of fever in these two cases was shorter than this average duration, indicating both Case 1 and Case 2 can get fever at the initial onset and can be controlled quickly. It is well known that fever can inhibit the viral replication and save response time for immune cells.

However, no fever or persistent fever often indicates the deficiency of immune system and a worsen prognosis, which also applies to patients with COVID-19^5^. These findings indicated the immune system in these two cases worked well which can be stimulated by the infection of virus^13^. Thus, we speculate rapidly controlled fever could be one of reasonable candidates to predict severity of patients in early stage. Together, the combination of the status of fever and level of lymphocytes may become an efficient indicator for early predication of prognosis in patients with COVID-19.

Plasma infusion treatment is a new therapeutic method for patients with COVID-19 and the efficiency is still needed to be investigated. In this case, the prolonged infection in Case 1 can be rapidly eliminated after infusion, implying this method might be a potent treatment for patients with COVID-19. However, the long-term outcomes and complications should be further investigated.

Although evidences collected from these two cases indicate a better prognosis, patients with long duration of viral shedding in mild type are likely to be neglected in crowd and may persist infecting surroundings and cause a new outbreak. The viral tests of Case 1 once changed into negative on illness days 47. However, it reversed into positive on illness days 49 (Fig. 1A). It is likely that the virus and the host got a dynamic balance. The viral load and the indicators of immune cells both maintained stable (Fig. 2). Although the viral duplication was suppressed, immune cells cannot clear out the virus. The Case 1 may tend to be a chronic infected case without infusion treatment, the virus and the host may even form a symbiotic relationship. We wondered how many patients involved in this situation. One important question is whether and how long this kind of patients keeps infective. The other important question is whether the “chronic infected patients” will infect through new route of transmission, such as sexual transmission. Moreover, due to the high mutation rate of retrovirus, we should keep close eyes on the status of this kind of patients and the infective ability.

We reported the clinical features of a special family cluster case, of which one member has the longest duration of viral shedding in current reports. Besides to the long duration, several points should also be concerned. Firstly, the virus infecting this family cluster case appeared low toxicity and low transmissibility. However, it also showed a prolonged infective ability and was hardly to be eliminated in body. This virus type may exist in human bodies for a long time and may be a risk to contribute to periodic outbreak in future considering the high mutation in COVID-19. Secondly, a rapidly controlled fever with normal level of immune cells may be a good predictor to identify patients with superior outcomes in early stage. This conclusion is needed to be validated in carefully designed clinical studies. Thirdly, plasma infusion could be an efficient therapy for patients with COVID-19. Last, long term health status and the infective ability of these people are needed to be followed up due to prolonged infection. Our report provides valuable information for public health management and for the decision making in triage and long-term follow-up and points out plasma infusion is a permissive treatment with superior efficiency.

## Data Availability

All the raw data can be provided by the corresponding authors upon reasonable request.

## AUTHOR AFFILIATIONS

From the Department of Disease Control and Prevention (L.T., T.Y., Q.W.), Department of Infectious Disease (B.Z.) and Department of Transfusion (S.Z.) in General Hospital of Central Theater Command, Department of Biochemistry and Molecular Biology in Third Military Medical University (Army Medical University) (X.K., F.Y., H.M.) and Department of Laboratory in No. 967 Hospital of PLA (B.L.).

## CORRESPONDING AUTHORS

Hongming Miao

Qiongshu Wang

## CONFLICT OF INTERESTS

The authors declare no competing interests.

### ACKNOWLEDGEMENTS

We thank all the doctors, nurses, public health workers, and researchers for the braveness in fighting against SARS-CoV-2 and the efforts to save the life of COVID-19 patients. This work was supported in part by award numbers 81872028 and 81672693 (H.M.) from the National Natural Science Foundation of China, cstc2017jcyjBX0071 (H.M.) from the Foundation and Frontier Research Project of Chongqing and T04010019 (H.M.) from the Chongqing Youth Top Talent Project.

## CONTRIBUTIONS

Li Tan and Xia Kang contributed equally to this work. Li Tan and Qiongshu Wang were responsible for the collection and summary of clinical data. Xia Kang and Hongming Miao wrote the manuscript. Hongming Miao was responsible for data integrity and manuscript submission.

---

**Fig. S1.**
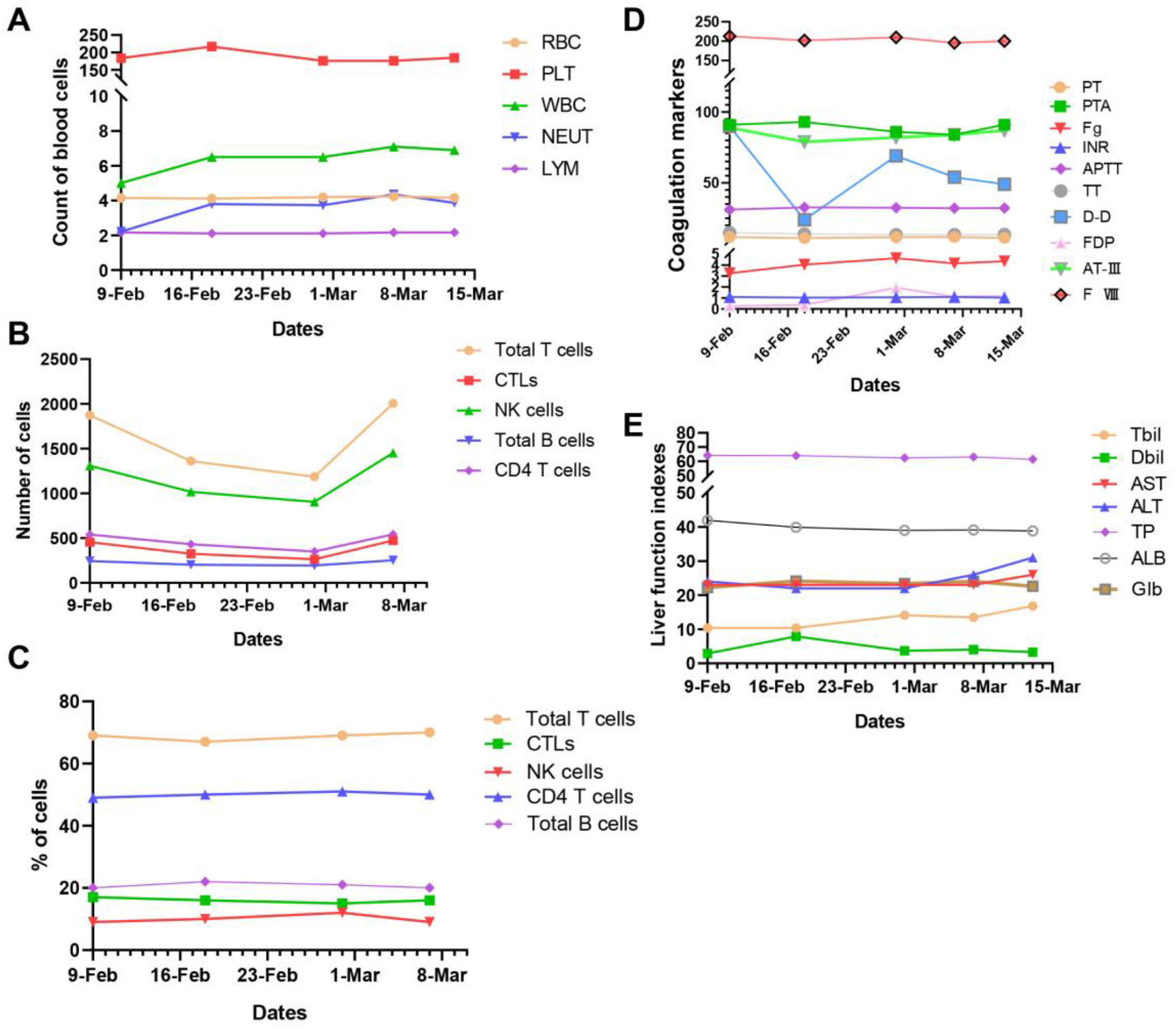
Dynamic tests of blood indexes in Case 1 after admission. (A) The count of blood cells. RBC (×10^12^/L), PLT (×10^9^/L), WBC (×10^9^/L), NEUT (×10^9^/L) and LYM (×10^9^/L) (B) The count of peripheral lymphocytes and NK cells. Total T cells (/ul), CTLs (/ul), NK cells (/ul),Total B cells (/ul) and CD4 T cells (/ul) (C) The percentages of peripheral lymphocytes and NK cells. (D) The tests for coagulation indicators. PT (s), PTA (%), Fg (g/L), APTT (s), TT (s), D-D (ng/ml), FDP (μg/ml), AT-III (%), F VIII (%) (E) The tests of liver function indicators. Tbil (μmol/L), Dbil (μmol/L), AST (U/L), ALT (U/L), TP (g/L), ALB (g/L), GLB (g/L) RBC, red blood cells; PLT, platelets; WBC, white blood cells; NEUT, neutrophils; LYM, lymphocytes; CTLs, cytotoxic T lymphocytes; NK cells, natural killer cells; PT, prothrombin time; PTA, prothrombin time activity; Fg, fibrinogen; INR, international normalized ratio; APTT, activated partial thromboplastin time; TT, thrombin time; D-D, d-dimer; FDP, fibrin/fibrinogen degradation products; AT III, antithrombin III; Tbil, total bilirubin; Dbil, direct bilirubin; AST, aspartate transaminase; ALT, glutamic-pyruvic transaminase; TP, total protein; ALB, albumin; Glb, globulin

